# Pragmatic Randomized Trial Assessing the Impact of Digital Health Technology on Quality of Life in Patients With Congestive Heart Failure: Design and Rationale

**DOI:** 10.1101/2021.11.19.21266591

**Authors:** Angela M. Victoria-Castro, Melissa L. Martin, Yu Yamamoto, Tariq Ahmad, Tanima Arora, Frida Calderon, Nihar R. Desai, Brett Gerber, Kyoung A. Lee, Daniel Jacoby, Hannah Melchinger, Andrew Nguyen, Melissa M. Shaw, Michael Simonov, Alyssa Williams, Jason Weinstein, F. Perry Wilson

**Affiliations:** Clinical and Translational Research Accelerator (CTRA), Department of Medicine, Yale University School of Medicine, New Haven CT; Department of Medicine, Section of Cardiology, Yale University School of Medicine, New Haven CT; Department of Medicine, Section of Rheumatology, Allergy, and Immunology, Yale University School of Medicine, New Haven CT; Department of Medicine, Section of Nephrology, Yale University School of Medicine, New Haven CT

**Keywords:** Digital Health Technology, Congestive Heart Failure

## Abstract

Heart failure is a complex syndrome that contributes significantly to mortality and morbidity in the Unites States. Self-management is an ACC/AHA-recommended management tool for chronic conditions, however, those with congestive heart failure have historically poor compliance, low health literacy, and comorbidities that lead to reduced adherence to therapies and lifestyle modifications. Digital health technologies have the potential to enhance care and improve self-management. This manuscript describes the rationale and challenges of the design and implementation of a pragmatic randomized controlled trial to evaluate the efficacy of three digital health technologies in the management of congestive heart failure. Leveraging the use of a fully electronic enrollment and consent platform, the trial will randomize 200 patients across heart failure clinics in the Yale New Haven Health system to receive either usual care or one of three distinct digital technologies designed to promote self-management and provide critical data to clinicians. Our primary outcome will measure the change in quality of life as assessed by the Kansas City Cardiomyopathy Questionnaire (KCCQ) at 3 months. Initial recruitment efforts have highlighted the large digital divide in our population of interest. Assessing not only clinical outcomes, but patient usability and ease of clinical integration of digital technologies will prove beneficial in determining the feasibility and success of the integration of such technologies into the healthcare system. Future learnings will illustrate strategies to improve patient engagement with, and integration of, digital health technologies to enhance the patient-clinician relationship.

**Clinicaltrials.gov:** NCT04394754

## INTRODUCTION

Heart failure (HF), a complex syndrome resulting in impaired ventricular function, is a significant cause of morbidity, mortality, and hospitalization in the United States. With over half a million new cases diagnosed each year, prevalence is growing and improvement in patient outcomes has plateaued, with 30-day readmission rates of up to 25% and poor 5-year mortality rates of 50%.^1, 2^

Pharmacologic guidelines for the treatment of HF include the use of ACE inhibitors, angiotensin II receptor blockers (ARBs), aldosterone antagonists, and beta-blockers.^2, 3^ Recent updates also include the use of angiotensin-neprilysin inhibitors (ARNI) and sodium-glucose cotransporter-2 inhibitors (SGLT2i), particularly for HF patients with reduced ejection fraction (HFrEF).^4^ While clinical trials have shown current treatments may reduce all-cause and/or cardiovascular-related mortality, renal outcomes, and HF-related hospitalizations, the effectiveness of these interventions in the population at large is hampered by slow uptake and inadequate medication adherence.^5-9^

Patient self-management, recommended by ACC/AHA guidelines, is an effective management tool in chronic conditions.^3, 10, 11^ This may involve methods to improve medication adherence, practiced behavioral changes, and active engagement in symptom recognition and education. Interventions improving medication adherence have reduced readmission and mortality rates among the HF population.^12^ However, HF patients have historically poor self-management, even with specialist support and social networks.^13, 14^ Noncompliance with medication and lifestyle modifications is common among older HF patients and contributes to disease exacerbation.^15-17^

Lack of health literacy is a major source of noncompliance, shown to be an independent risk factor for both mortality and hospitalization.^18-20^ Additional comorbidities leading to high degrees of polypharmacy also contribute to nonadherence.^21-23^ Combined, this data highlights the need for novel strategies that improve HF patient engagement and self-care.

The use of digital health technologies has the potential to enhance and personalize care and improve the patient-clinician relationship.^24^ There are many potential benefits to both the patient and provider. Clinicians can gain access to inter-visit clinical data that may guide treatment and allow for earlier identification of clinical decompensation. Patients may experience improved health literacy, self-monitoring, and compliance, and become empowered to take a more active role in their care. Overall, digital health technology has the potential to enhance personalized care, increase clinical efficiency, and reduce resource utilization.

While digital health interventions are promising, challenges of clinical workflow, physician burden, and usability must be addressed.^25^ User-friendly platforms that both encourage patient engagement and provider adoption are essential to the success of digital interventions, highlighting the need for comprehensive randomized clinical trials that address the effects of digital health on clinical outcomes, patient usability, and clinical integration. ^26, 27^

Our pragmatic randomized controlled trial examines the effectiveness of three distinct digital health technologies versus usual care in the management of congestive heart failure (CHF): a “smart” scale (Bodyport), an automated conversational platform (Conversa), and a coaching application (Noom). The three technologies differ fundamentally in design, user interface, and data collected. However, the use of a common control group allows for the evaluation of multiple potential interventions in an efficient trial design space. Thus, this trial will provide insight into which types of technologies can be best integrated into clinical practice and patient experience.

The primary outcome will measure the change in quality of life as assessed by the Kansas City Cardiomyopathy Questionnaire (KCCQ), while secondary outcomes will assess clinical outcomes and efficiency as well as patient usability and perception.

We hypothesize that digital health interventions will improve quality of life of heart failure patients through one or both of two potential mechanisms. Digital health interventions that track symptoms and detect early warning signs of disease exacerbation which are then relayed to providers may result in medication and/or lifestyle adjustments that can mitigate symptoms and thus improve patient quality of life. Secondly, technologies that allow for self-tracking of symptoms and that provide education regarding a condition and appropriate self-care may result in enhanced patient empowerment, medication adherence, and increased involvement of the patient in their own care management. Consistent encouragement and self-care prompts may lead patients to make better daily lifestyle choices and more frequently communicate concerns with providers, both of which may lead to fewer symptoms and thus improved quality of life.

## METHODS AND ANALYSIS

This pragmatic study is an unblinded, 4-arm, parallel-group, randomized, controlled trial to determine effectiveness of three digital health technologies versus usual care in improving quality of life for patients with congestive heart failure. This study will assess clinical outcomes, clinical efficiency and burden, and patient usability and perception. The trial is conducted under approval of the Yale University Institutional Review Board and adheres to the principles of the Declaration of Helsinki. The trial is pre-registered at clinicaltrials.gov (NCT04394754)

### Participants

This study will enroll patients actively managed by general heart failure clinics located throughout the Yale-New Haven Health system that follow patients closely for optimization of care and capture a socioeconomically diverse patient population.

Patients must have a current diagnosis of congestive heart failure without regard to ejection fraction. Eligibility is restricted to those 18-79 years of age, as the pediatric population requires unique management and elderly patients may not benefit from digital intervention due to lack of digital literacy or advanced disease.^28^

### Exclusion criteria

This study will exclude patients with an advanced stage of HF or comorbidities that might limit usability or benefit of the devices. To this end, patients with class IV HF – as determined by the treating provider, who have received a heart transplant, have a ventricular assist device, are diagnosed with stage 4 or end stage renal disease (or eGFR <30), are in hospice, are diagnosed with dementia, are determined by their physician to have a life expectancy of <6 months, are pregnant, or those otherwise unable to consent are excluded. Those in unstable living situations, or who are incarcerated, are excluded due to potential difficulties with follow-up and the potential for shared use of devices, which is discouraged in this study. Patients currently enrolled in another digital health study will be excluded to avoid potentially contaminating effects. Finally, we will exclude patients who weigh over 400 pounds or are unable to stand for 30 seconds unassisted to ensure that all patients can use the Bodyport scale should they be randomized to this group.

We are careful not to exclude patients on the basis of a lack of technological access. Patients will be given a prepaid smartphone with a limited data plan if they otherwise have no access to one and will be guided through email account creation if needed, as an email address is required to receive study-related communications.

### Interventions

Upon enrollment, patients will be randomized to one of four study arms, three of which involve the receipt of a digital health technology (see below). Patients will be encouraged, but not obligated, to engage with their technology daily. We were careful not to mandate protocols regarding frequency of use, such that we can determine real-world use and applicability. We will emphasize to participants that no digital health technology in this study should replace usual care and will advise participants to follow the care plan outlined by their primary provider and consult with their physician or 911 during emergencies.

The three technologies included in this study were chosen through a vendor assessment process, which started with an evaluation of the market landscape for digital solutions that met essential needs of patients with CHF. A competencies matrix with weighed criteria was developed and served as the basis for an initial request for information to different companies. This information included questions to inform company study operations, evidence and outcomes, patient safety, regulatory, quality, and technical requirements. Furthermore, a supplemental proof and demonstration of outcomes in CHF was requested, narrowing the final list of candidate digital solutions to 10. Bodyport, Conversa and Noom were selected as the interventions in this study after an in-person showcase where all 10 solutions were evaluated by the study coordination team.

### Bodyport smart scale

Bodyport is a data-driven smart scale that measures clinically relevant markers of fluid and cardiovascular status, enabling an integrated, longitudinal assessment of congestion and perfusion to help guide personalized heart failure management.^29^ Measurement data is uploaded over a cellular connection to a clinician dashboard (outside the EHR) for review by both the study and clinical teams **(Figure 1)**. Patients can access a similar dashboard, the Bodyport Patient Portal, to track measurements and view learning modules related to heart failure and effective self-management. These modules consist of videos with transcripts, and learning checks composed by a set of multiple-choice questions at the end. The content was created by Bodyport and is based on AHA/ESC recommendations for Heart Failure education. For this study, only a subset of Bodyport biomarkers was used by the study team: body weight, impedance – a marker of fluid overload, and heart rate.

**Figure 1:**
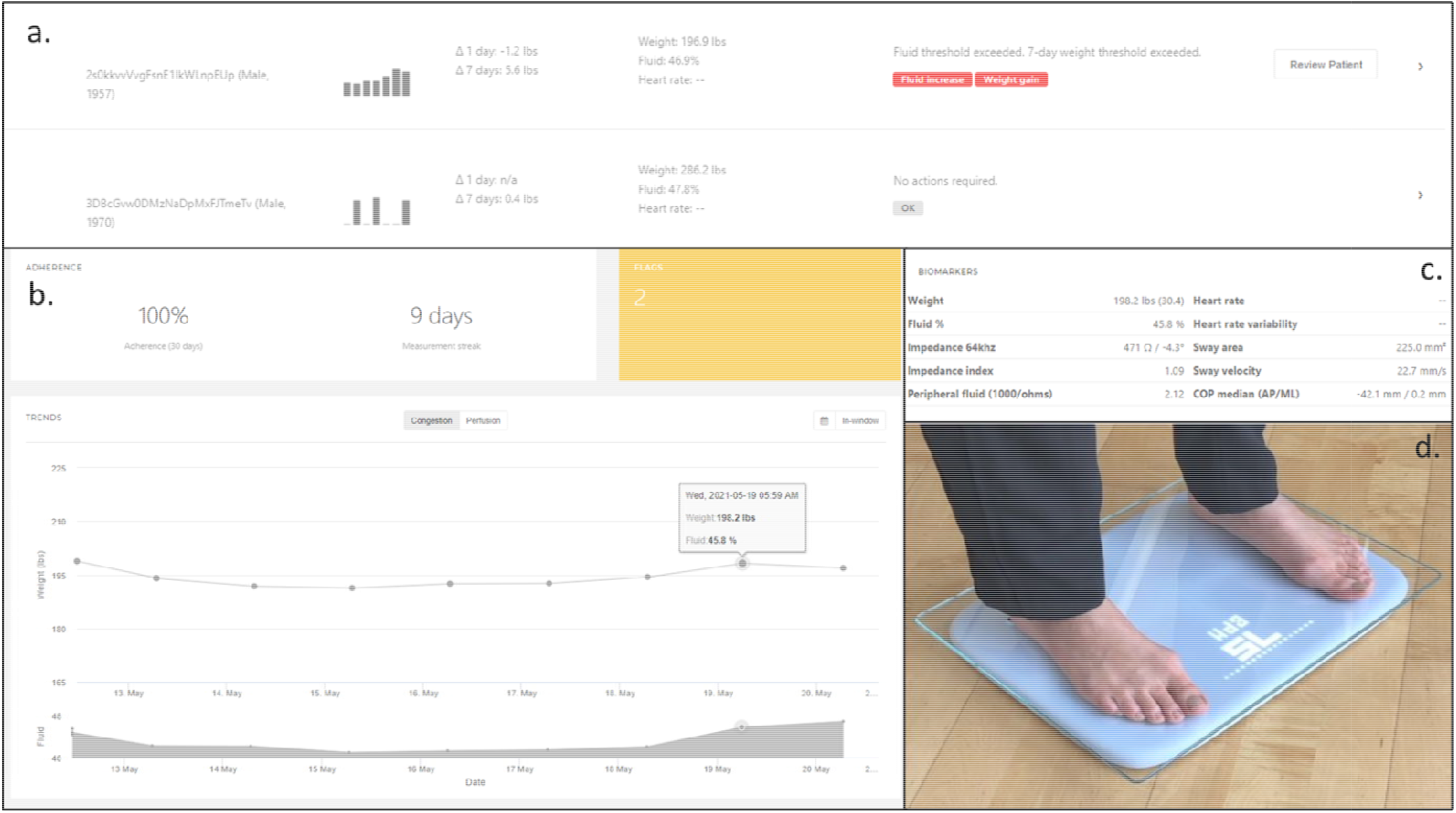
Important elements of the Bodyport physician-facing dashboard. **a**. Global dashboard displaying overall trends and alerts. **b**. Detailed individual patient data showing trends in weight and perfusion. **c**. Advanced metrics of cardiovascular function. **d**. Representative Bodyport scale.

### Conversa

Conversa is an automated conversational platform designed to engage patients in self-management. In this study, patients participate in a heart failure-oriented program, where they may complete educational modules and engage in regular brief (approximately 5 minutes) “clinically intelligent” chats with pre-scripted responses that cardiovascular physicians have vetted to ensure appropriateness and applicability to the study population. The chats are designed to collect patient-generated health data regarding overall health, symptoms, and self-care, which is uploaded to a clinician dashboard that can be viewed by both the study and clinical teams **(Figure 2)**.

**Figure 2:**
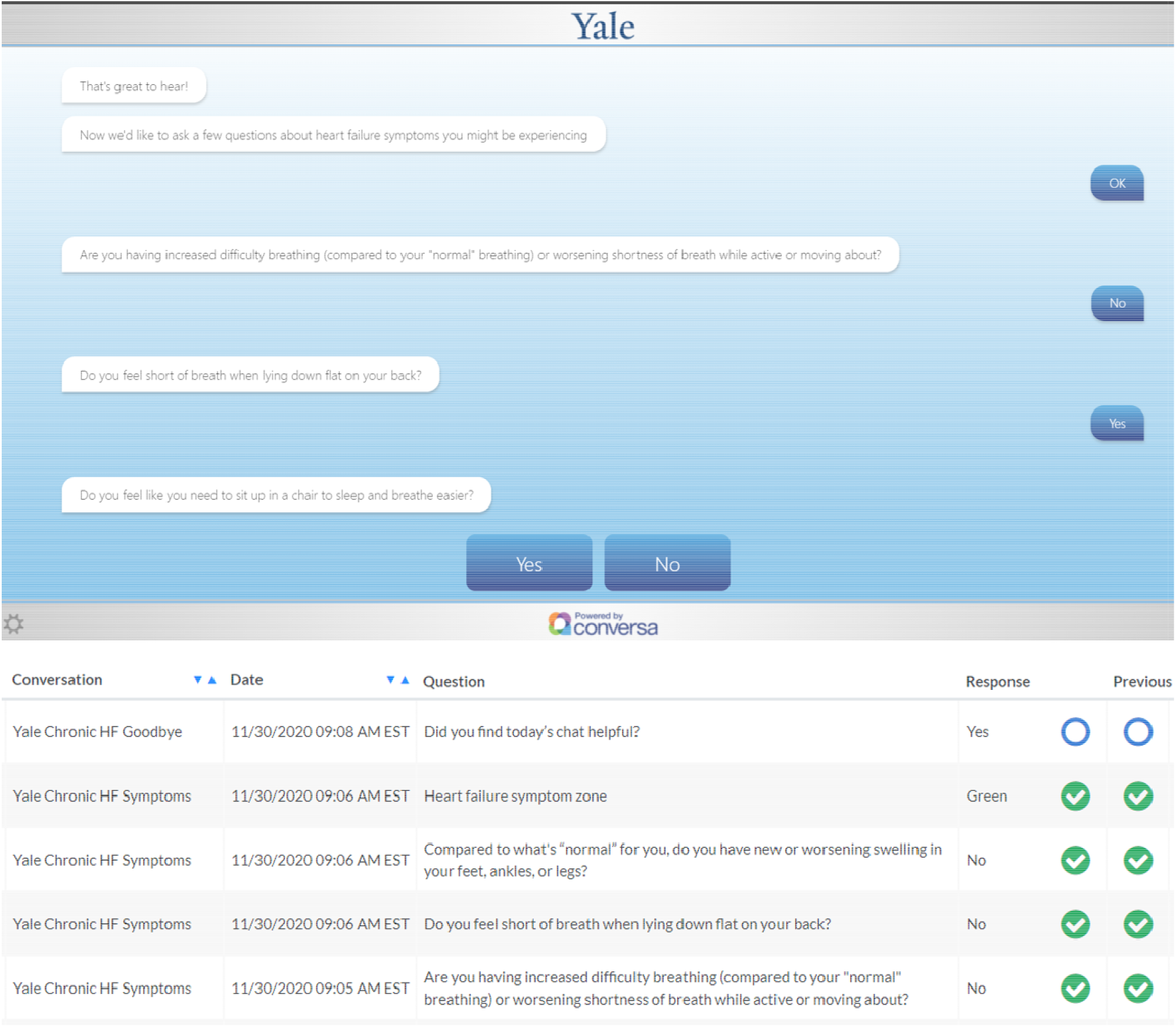
**Top Panel:** Example “chat” for a heart failure educational module. **Bottom Panel:** Example of the physician-facing Conversa dashboard with a view of patient responses over time.

### Noom

Noom is a data-driven smart phone coaching application that uses evidence-based behavior change techniques to coach patients towards better self-management of their syndrome.^30^ On this heart failure-tailored program, patients can track medication adherence, physical activity, symptoms, self-reported physiological parameters (such as blood pressure and glucose levels) and eating habits via food and macronutrient logging. Live coaching support **(Figure 3)** and the possibility of interaction with other users through support groups within the application is also available. The Noom coaches are trained during three months on Noom’s technology and approach which is based on motivational interviewing and Cognitive Behavioral-Therapy, and they are certified as lifestyle coaches by the National Board for Health and Wellness Coaching. Clinically significant changes based on patient-reported data will be communicated to the team via a Noom coach.

**Figure 3:**
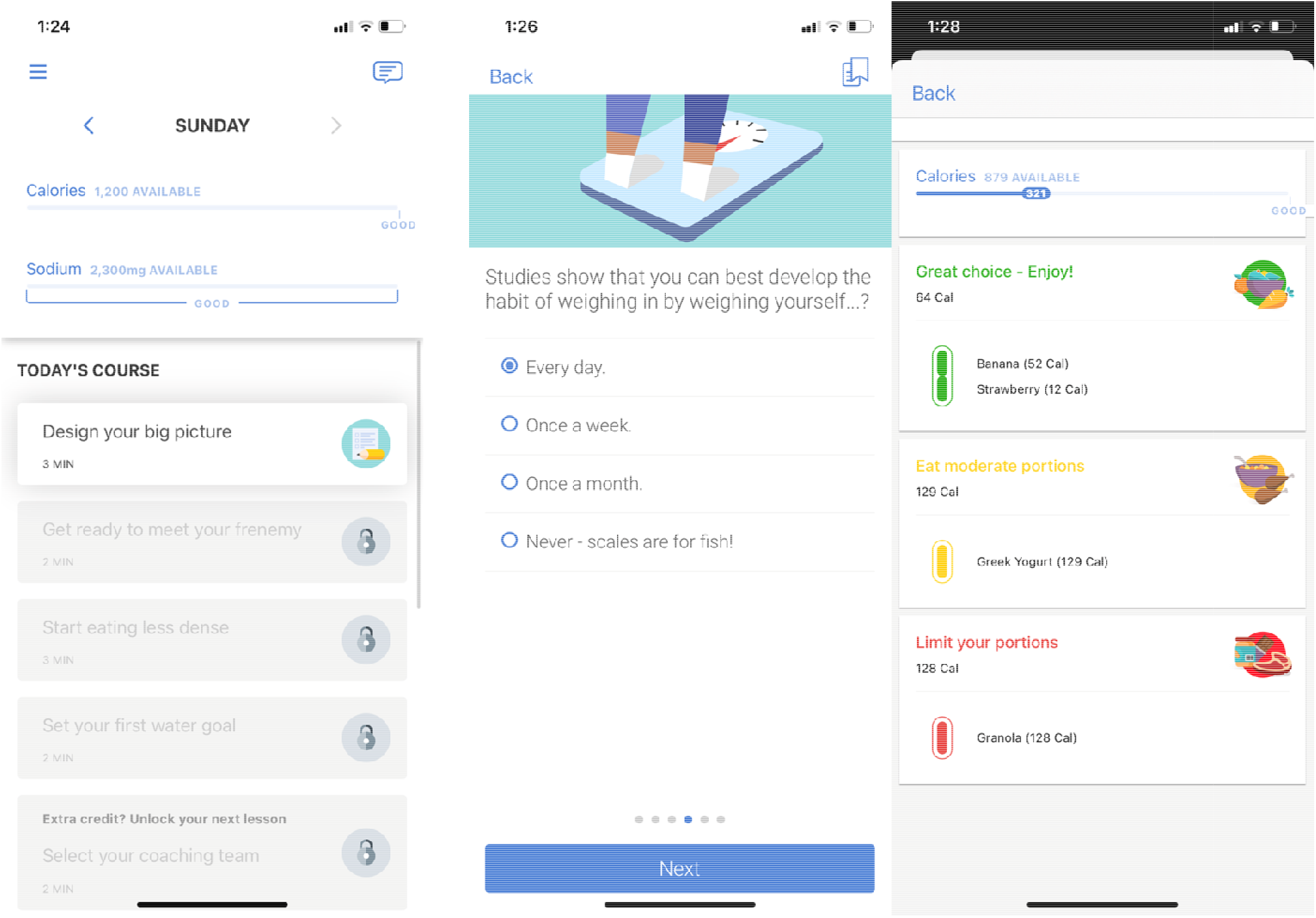
Representative Noom program displaying daily program activities, educational programming, and nutrient logging.

### Usual Care

Patients randomized to usual care will not receive a digital health device. They will be asked to continue with their usual care as prescribed by their health care team. Patients will still be asked to complete all study-related surveys and will be followed similarly as those in the intervention groups.

### Recruitment

Eligible patients will be prospectively identified via electronic chart review. Those who meet all study criteria will either be approached during a clinic visit or contacted via phone calls, text messaging, or electronic health record communication. Patients will be given a thorough explanation of the study and have any questions answered.

Interested patients will be guided through a web-based enrollment platform that collects eligibility and demographic information, allows patients to read and sign an electronic consent, and directs the patient to create a study participant profile. Once this profile is complete, the patient is enrolled and is directed to their baseline surveys. All study enrollment and survey completion is performed in the VARA™ platform (Medullan)^31^.

### Randomization

Upon completion of enrollment and all baseline surveys, patients will immediately receive their assignment and any further instruction for receiving and activating any digital health technologies. Randomization is achieved through a permutated block randomization scheme, stratified by clinic, created by an independent statistician. This will ensure that case-mix differences across study sites will not drive observed differences in technology effectiveness. Cluster randomization was not considered given the small number of clusterable entities, the heterogeneity among the clinics, which may introduce confounders, and the low risk of contamination across study arms. Randomization at the provider level was infeasible given the small number of clinicians providing care at each clinic. Additionally, patients may be cared for by multiple providers across clinic visits.

### Blinding

This is an open label study, as the study team will be actively monitoring incoming data from participant devices to identify and properly triage red flag warnings (described below). Clinicians may also access dashboards and receive red flag warnings regarding their patients.

### Patient Experience and Follow-Up

The patient experience timeline is outlined in **Figure 4**. All participants will be followed for six months. During three remote study-related visits at enrollment, at 3 months and 6 months, patients complete a set of surveys, including the Kansas City Cardiomyopathy Questionnaire (KCCQ), a medication adherence survey (PROMIS Med Adherence Scale)^32^, a health literacy assessment (Brief Health Literacy Screen, BHLS)^33^, and a technology literacy assessment (developed in-house, see supplemental material). The information will be used to ascertain baseline data and track progress throughout the study in each domain.

**Figure 4:**
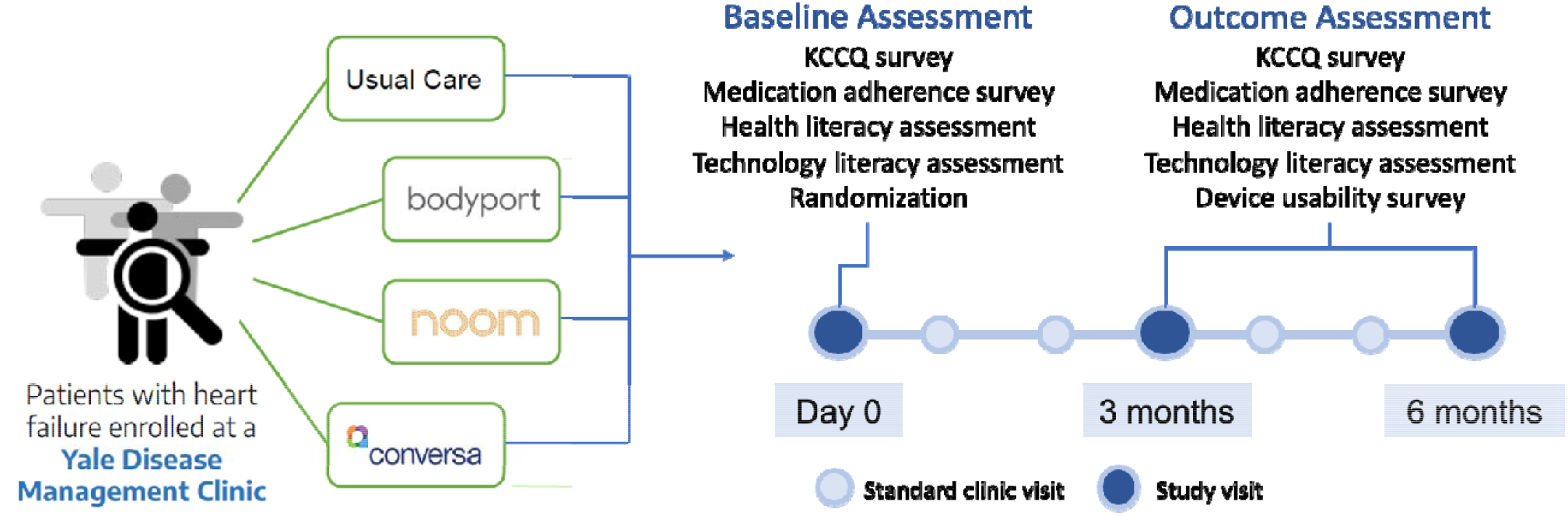
Central illustration. Patients are recruited from heart failure clinics and randomized to one of three digital health interventions or usual care. They are then followed for six months with both clinic and telephone visits. The primary outcome is change in the Kansas City Cardiology Questionnaire (KCCQ) at 3 months.

Throughout the study, participants will be encouraged, though not mandated, to use assigned technology regularly, and will continue to attend clinic visits per usual at their provider’s discretion. Patient- and device-generated health and usage data will be passively collected for outcome determination. During follow up visits, clinical data for secondary outcome determination will be collected in addition to survey completion, and those randomized to an intervention arm will also be directed to complete a survey assessing usability and satisfaction with the technology.

### Clinician Experience

Conversa and Bodyport data are uploaded to EHR-independent dashboards visible to both study and clinical teams. No standardized protocol exists regarding clinician access or use of this data so as to minimize clinician burden, with the exception of device triggers, explained below. We will emphasize to clinicians that study-related data is supplementary to regular care and should only be used in the context of all available medical information.

### Red Flag Warnings

We have pre-specified several device-specific “triggers” that will appear as red flag warnings in each of the technology platforms when a particular data point reaches a pre-determined threshold **(Table 1)**. For Bodyport, the research team accesses an online dashboard twice a day to identify red flag warnings; for Conversa, we receive emails when there is a trigger that needs to be revised in their online dashboard; and for Noom, each of the coaches email us directly to discuss the specific red flag warnings of their participants. Each trigger is vetted for clinical applicability by cardiovascular physicians on the study team and will be communicated to the clinical team via messages through the EHR platform. The patient care team will be asked, but not mandated, to follow up with the patient via a brief “check-in” call and report any new HF symptoms and whether the patient’s care plan has changed as a result. Responses are encouraged to demonstrate how these technologies alter or improve patient care. While we recognize that specific recommendations or algorithms in conjunction with a red flag warning may be useful in eliciting changes in care by providers, our goal was to minimize clinician burden in the context of a high alert environment where alert fatigue is possible. Further, in this pilot study, we aimed to keep minimal risk to patients while assessing general efficacy and acceptance of different technologies that can be explored more deeply in future work. Thus, in the current study we provide no additional standardized protocols beyond these “check-in” calls and leave further care to the clinical team’s discretion. Additionally, should frequent alerts burden clinicians, our protocol gives flexibility to modify triggers for greatest clinical utility.

**Table 1:** Technology-specific triggers that generate red flag warnings. SPB = systolic blood pressure; DBP = diastolic blood pressure; \**Calculates the difference between the in-window measurement today and the lowest in-window measurement over the past 24 hours or 7 days. In-window is 4-10am to minimize contamination by daily fluctuations in weight. Impedance, used as a marker for lower extremity edema, calculates changes in the in-window minimum and maximum values over a 5-day period*.

| Technology | Trigger |
| --- | --- |
| Bodyport | Increase in 3lbs over 24 hours*<br>Increase in 5lbs over 7 days*<br>Decrease in impedance of 30% over 5 days<br>HR less than 60 bpm sustained over two days<br>HR greater than 100 bpm sustained over two days |
| Conversa | Answer "Yes" to having shortness of breath that won't go away<br>Answer "Yes" to having to sit up to sleep and breathe easier<br>Answer "Yes" to increased difficulty completing daily activities<br>Answer "Yes" to having concerning symptoms (sudden chest pain, discomfort while resting)<br>Inputs a SBP >180 mmHg<br>Inputs a SBP <90 mmHg<br>Inputs a DBP >120 mmHg<br>Inputs a DBP <50 mmHg |
| Noom | Inputs a SBP >170 mmHg<br>Inputs a SBP <90 mmHg<br>Increase in 3lbs over 24 hours<br>Increase in 5lbs over 7 days |

### Primary Outcome

Our primary outcome is change in quality of life at three months as measured by the KCCQ, which has shown to be responsive and sensitive at this time point.^34^ This assessment is a standardized 23-item questionnaire, validated in multiple heart failure populations, that quantifies the physical limitations, frequency of symptoms, self-efficacy, social interference, and quality of life associated with living with HF. Scores range from 0-100, with higher scores indicating a more favorable status.

### Secondary Outcomes

Secondary outcomes of interest are listed in **Table 2**. Three categories of outcomes were selected as measures of successful digital health products: (1) clinical outcomes (2) clinical efficiency and (3) usage metrics indicating patient usability, ease of use, understanding of the assigned device and perception of engagement with health condition after the usage of technology. Each metric will be assessed at the three-month and six-month time point to determine the stability of the effect of digital health technology over time as clinic visits may decrease or as HF care is optimized. KCCQ scores will be reassessed at six months.

**Table 2:** Secondary Outcomes

| Secondary Outcomes |  |
| --- | --- |
| Clinical Outcomes | Hospital admissions<br>Rates of guideline directed medical therapy<br>ED visits<br>Mortality<br>Clinic no-show rates<br>AKD development |
| Patient Usability | Satisfaction with device/technology<br>Frequency of use (overall and feature level)<br>Content rating<br>Perceived impact |
| Clinical Efficiency | Amount of intravisit contact<br>Number of clinic visits<br>Provider assessment of global disease severity<br>Effect on workload and efficiency |
| Usage metrics | Number of enrolled patients<br>Number of incomplete enrollments<br>Number of complete and incomplete assessments<br>Time to complete initial assessments<br>Number of complete and incomplete product set-ups<br>Number of products that completed initial data sync<br>Percent of patients with last screen assessed in session<br>Number of active (>1 login per week) patients per week<br>Average number of logins per week<br>Statistic for each session (time of day, day of week, number of page views, origin of login (direct, email))<br>Average number of interactions<br>Average number of daily check-ins<br>Number of patients not using product per week<br>Time of day of product usage<br>Completeness of data provided per question in session<br>Dwell time per feature |

### Statistical Analysis

Baseline comparisons across the study arms will be conducted with chi-square tests for categorical variables, applying the Maentel-Haenszel correction for stratification by clinic site. Continuous variables will be compared using the Van-Elteren test for continuous, non-parametric variables and stratified t-tests for normally distributed variables.

The primary analysis will be a mixed effects model comparing change in KCCQ over time in the intervention group versus the control, without accounting for a center-by-treatment interaction. In secondary analyses, we will model that interaction to determine if certain interventions are uniquely effective in certain centers.

Secondary and exploratory analyses will examine the outcomes listed above using the same procedure applied to baseline variables for outcomes assessed at one time point and using mixed-effects models for continuous outcomes assessed at multiple time points. We will also perform an analysis of the effect of digital health interventions in general versus usual care by combining the three digital health interventions into a single analytic group.

### Power Calculations

The study was designed to provide adequate power to detect a clinically significant change in the primary outcome measure, the Kansas City Cardiomyopathy Questionnaire score. The KCCQ composite score ranges from 0 – 100 (with 100 being the best possible score), and a change of 10 points has been determined to be the minimum clinically important difference. Within a given patient, the score is relatively stable in the absence of clinical changes, with a within-patient standard deviation of 11.8 for patients with no change in clinical status.

To detect the minimum clinically important difference of a 10-point change in the KCCQ, each arm of the trial must include 40 patients across all centers for 90% power at an alpha threshold of 0.017, giving a total of enrollment of 160. Given uncertainties around the intraclass correlation coefficient (assumed to be 0.10 here and reflecting each clinic’s overall efficacy) the target sample size is inflated to 200, or 50 per trial arm. The lower alpha threshold represents a Bonferroni correction that provides statistical power to test multiple comparisons – each of the three digital health interventions against the control intervention. We will attempt to balance recruitment across the centers with a per-center target of five individuals per trial arm.

### Interim Analysis

We will have one interim analysis at the mid-point of the trial (50% enrollment). This will allow us to alter the sample size or stop the trial earlier for ethical considerations, unexpected adverse events, or high efficacy. The trial will stop for declaring efficacy if the effect size is large. We will stop the trial for efficacy for a p-value threshold of < 0.0006. Given the interim analysis, the threshold p-value for statistical significance at the end of the trial will be 0.016.

### Preliminary Findings: Recruitment Challenges

At the time of manuscript submission, we have successfully recruited 182 patients and are conducting follow-up visits and data collection, illustrating the feasibility of our proposed methodology. While feasible, recruitment of the population of interest was not without its challenges, which were overcome through a series of learnings and adaptations that may prove useful for future studies.

This study was designed to be fully remote, with an online enrollment platform and digital technologies that patients can use from home. A remote design has obvious advantages in today’s current climate amongst a global pandemic, when remote doctor’s visits and social distancing have become the norm and appeals to patients who already feel burdened by many in-person clinic visits.

This platform was carefully designed for ease of use and was put through an extensive user acceptance testing period to identify flaws. Despite its benefits, a fully remote design has revealed the large technological gap our target population experiences, presenting a challenge for those less technologically-savvy. Eligible patients are often elderly and reside in areas of lower socioeconomic status, both demographics that are affected by the digital divide.^35, 36^ The development of a mobile- and user-friendly platform proves essential in reducing these enrollment barriers in this and future studies. Further, some patients are reluctant to provide personal information or click links sent by unknown coordinators. Many are simply difficult to reach by phone. Our study design has undergone several adaptations to enhance user experience and improve remote recruitment. Multiple adjustments were made to the enrollment website to reduce user error and improve website navigation, and coordinators were available to fully guide participants through the online enrollment process, completing questions on behalf of the participant via the phone if necessary. The use of pre-scripted text messages and MyChart communication were used to make patients primed for receiving study-related recruitment calls. Further, use of a HIPAA-compliant dialer was used for customization of caller ID to a local area code.

While we recognize that, because of our exclusion of those patients over 80 years of age, this study may be intrinsically biased towards those with more digital literacy, we hope that the digital divide we have experienced with this slightly younger population of heart failure patients and the subsequent adjustments and learnings will guide future work with digital health technologies in similar, and perhaps older, populations.

## DISCUSSION

Digital health technology has proven an effective modifier of patient behavior in conditions such as mental illness, diabetes, and atrial fibrillation.^37-40^ Few observational studies and small randomized trials have assessed remote monitoring systems and wearable devices in the HF population.^41-43^ Our goal is to evaluate the efficacy of three non-invasive and distinct technologies that may directly affect patient quality of life, behavior, and engagement with their providers.

For any digital technology meant to affect patient behavior, user experience is a key design– despite collecting useful data, digital technology provides little value unless used regularly by patients or clinicians. Historically, patient retention rates of digital health technologies are low due to a lack of user-centered design processes.^27, 44^ Within the HF domain, it is particularly essential to deliver functional capabilities that cater to an elderly population.^45^ Among seniors, user rates of everyday technology are drastically lower than that of the general population, with only modest yearly increases.^28^ While willingness to use technology has increased among the elderly, lack of familiarity and the need for facilitated training may create barriers.^46, 47^ Whether this digital gap will translate into low rates of device usage and lack of effect on clinical outcomes, particularly in the context of a remotely designed study, remains to be determined.

The use of digital health technologies in the healthcare space has the potential to improve overall patient care and strengthen the provider-patient relationship. Improvement of patient engagement and health literacy may lead to better self-care habits and treatment compliance, while technology may allow clinicians to better identify disease exacerbation and better personalize care. A variety of primary and secondary study endpoints will address three major contributors to the success of any digital health technology: usability, clinical integration, and effect on patient outcomes. Further learnings will illustrate strategies to better engage a less technologically-savvy population of HF patients, ensuring that this patient group is not excluded from potentially beneficial digital health solutions.

## Data Availability

All data produced in the present work are contained in the manuscript

## Abbreviations

HF: heart failure
HFrEF: heart failure with reduced ejection fraction
HFpEF: heart failure with preserved ejection fraction
CHF: congestive heart failure
KCCQ: Kansas City Cardiomyopathy Questionnaire

## Notes

**Declaration of interests:** This study is funded by Boehringer Ingelheim. However, the authors have full jurisdiction over all aspects of study design, conduct, analysis, and publication.

FPW: Founder of Efference, LLC, a medical communications company. Receives support from NIH R01DK113191, R01HS027626, P30DK079310

NRD: Under contract with the Centers for Medicare and Medicaid Services to develop and maintain performance measures used for public reporting and pay for performance programs. Reports research grants and consulting for Amgen, Astra Zeneca, Boehringer Ingelheim, Cytokinetics, MyoKardia, Relypsa, Novartis, and SCPharmaceuticals.

### Competing Interest Statement

This study is funded by Boehringer Ingelheim. However, the authors have full jurisdiction over all aspects of study design, conduct, analysis and publication.
Dr. Wilson is Founder of Efference, LLC, a medical communications company. Receives support from NIH R01DK113191, R01HS027626, P30DK079310.
Dr. Desai is under contract with the Centers for Medicare and Medicaid Services to develop and maintain performance measures used for public reporting and pay for performance programs. Reports research grants and consulting for Amgen, Astra Zeneca, Boehringer Ingelheim, Cytokinetics, MyoKardia, Relypsa, Novartis, and SCPharmaceuticals.

### Clinical Trial

NCT04394754

### Funding Statement

This study is funded by Boehringer Ingelheim. However, the authors have full jurisdiction over all aspects of study design, conduct, analysis and publication.

### Author Declarations

The trial is conducted under approval of the Yale University Institutional Review Board and adheres to the principles of the Declaration of Helsinki.

